# Clinical Course And Risk Factors For In-hospital Death In Critical COVID-19 In Wuhan, China

**DOI:** 10.1101/2020.09.26.20189522

**Authors:** Fei Li, Yue Cai, Chao Gao, Lei Zhou, Renjuan Chen, Kan Zhang, Weiqin Li, Ruining Zhang, Xijing Zhang, Duolao Wang, Yi Liu, Ling Tao

**Affiliations:** Department of Cardiology, Xijing Hospital, Fourth Military Medical University, Xi’an, China; Huoshenshan Hospital, Wuhan, China; Clinical Laboratory, Xijing Hospital, Fourth Military Medical University, Xi’an, China; Department of Critical Care Medicine, Jinling Hospital, Nanjing, China; Surgical ICU, Department of Anesthesiology, Xijing Hospital, Fourth Military Medical University, Xi’an, China; Department of Clinical Sciences Liverpool School of Tropical Medicine Pembroke, Liverpool, United Kingdom

## Abstract

**BACKGROUND:** The risk factors for mortality of COVID-19 classified as critical type have not been well described.

**OBJECTIVES:** This study aimed to described the clinical outcomes and further explored risk factors of in-hospital death for COVID-19 classified as critical type.

**METHODS:** This was a single-center retrospective cohort study. From February 5, 2020 to March 4, 2020, 98 consecutive patients classified as critical COVID-19 were included in Huo Shen Shan Hospital. The final date of follow-up was March 29, 2020. The primary outcome was all-cause mortality during hospitalization. Multivariable Cox regression model was used to explore the risk factors associated with in-hospital death.

**RESULTS:** Of the 98 patients, 43 (43.9%) died in hospital, 37(37.8%) discharged, and 18(18.4%) remained in hospital. The mean age was 68.5 (63, 75) years, and 57 (58.2%) were female. The most common comorbidity was hypertension (68.4%), followed by diabetes (17.3%), angina pectoris (13.3%). Except the sex (Female: 68.8% vs 49.1%, P=0.039) and angina pectoris (20.9% vs 7.3%, *P* = 0.048), there was no difference in other demographics and comorbidities between non-survivor and survivor groups. The proportion of elevated alanine aminotransferase, creatinine, D-dimer and cardiac injury marker were 59.4%, 35.7%, 87.5% and 42.9%, respectively, and all showed the significant difference between two groups. The acute cardiac injury, acute kidney injury (AKI), and acute respiratory distress syndrome (ARDS) were observed in 42.9%, 27.8% and 26.5% of the patients. Compared with survivor group, non-survivor group had more acute cardiac injury (79.1% vs 14.5%, *P*<0.0001), AKI (50.0% vs 10.9%, *P*<0.0001), and ARDS (37.2% vs 18.2%, P=0.034). Multivariable Cox regression showed increasing hazard ratio of in-hospital death associated with acute cardiac injury (HR, 6.57 [95% CI, 1.89-22.79]) and AKI (HR, 2.60 [95% CI, 1.15-5.86]).

**CONCLUSIONS:** COVID-19 classified as critical type had a high prevalence of acute cardiac and kidney injury, which were associated with a higher risk of in-hospital mortality.

## INTRODUCTION

Beginning in December 2019, the COVID-19 has caused an international outbreak of respiratory illness. By March 30, 2020, the confirmed COVID-19 patients had exceeded 700 thousand. The soaring of COVID-19 has been seen as one of the most serious hazards to global health.

COVID-19 is clinically classified as four types: mild, moderate, severe and critical. Critical patients have critical pulmonary injury, systemic inflammatory status and a very high mortality(1-3), which leads to tremendous difference in clinical course, medical intervention and prognosis compared with mild to severe type. Illustration of demographics, clinical characteristics, complications and treatment outcome of critical patients is practically important to get further insights into the early origins of adverse outcomes and may ultimately be relevant for developing clinical prediction models. Although some COVID-19 case series and studies have been reported previously(4-6), to our knowledge, there are limited studies only including critical patients and specifically focusing on adverse outcomes and the predictive factors.

In the present study, we retrospectively included 98 consecutive patients with critical COVID-19 in Huoshenshan hospital (Wuhan, China). We described the patient demographics, laboratory findings, treatment & complications and further explored risk factors of in-hospital death for these patients.

## METHODS

### Study design and Participants

This is a single-center, retrospective cohort study. A total of 2074 consecutive patients with COVID-19 were screened in Huo Shen Shan Hospital from February 5, 2020 to March 4, 2020. Huo Shen Shan Hospital was opened since February 3, 2020, designated by the government only for treating COVID-19. After excluding the mild, moderate and severe patient, 98 patients classified as critical type on or after admission were included in the final analysis. The final follow-up date was March 29, 2020. Patients with COVID-19 enrolled in this study were diagnosed according to World Health Organization interim guidance (7). The classification of COVID-19 is according to the COVID-19 diagnosis and treatment program issued by the National Health Commission of China(8): (1) Mild type: mild symptoms, and no pneumonia imaging showed. (2) Moderate type: patients with fever, respiratory symptoms, and/or pneumonia imaging. (3) Severe type: patients with any of the following: shortness of breath, respiratory rate (RR)>30 times/ min; oxygen saturation <93% in resting state; Arterial partial pressure of oxygen / Fraction of inspired oxygen (PaO_2_/ FiO_2_) <300 mmHg (1mmHg=0.133 kPa) and the pulmonary imaging showed more than 50% lesion progression within 24 ~ 48h; (4) Critical type: patients with any of the following conditions: respiratory failure requiring mechanic ventilation, shock, or organ failure requiring intensive care. Patients were divided into Survivor Group or Non-survivor Group according to their clinical outcome.

This study was approved by the National Health Commission of China and the institutional review board at Huoshenshan Hospital (HSSLL025). Written informed consent was waived by the Ethics Commission of the designated hospital for patients with emerging infectious diseases.

### Data Collection

The demographics, laboratory findings, treatment & complications for participants during hospitalization were collected from electronic medical records by 2 investigators. All data were independently reviewed and entered into the computer database by 2 analysts.

### Clinical Endpoints

The primary endpoint was all-cause mortality during hospitalization. Other endpoints included: 1) Cardiac injury, defined as blood levels of cardiac injury markers (hs-TNI or CK-MB) above the upper reference limit.; 2)Acute respiratory distress syndrome (ARDS), defined according to the Berlin definition(9); 3)Acute kidney injury, identified according to the Kidney Disease: Improving Global Outcomes definition (KDIGO)(10).

### Statistical Analysis

Continuous data are presented as mean SD or median (interquartile range) and compared using the Student’s t-test or the Mann-Whitney test depending on their distributions. Categorical variables were expressed as frequencies with percentages and compared with the Chi-square or Fisher exact tests as required. Kaplan-Meier curves were constructed to estimate the cumulative incidence of death and were compared using the log-rank test. Cox proportional hazards models were used to identify risk factors for the occurrence of death. Results are reported as HRs and 95% CIs. Only covariates associated with the risk of death at a univariate analysis with P <0.1 were then included in the multivariate analysis of in-hospital death. All statistical tests were two-tailed, and P values were statistically significant at <0.05. All data were analyzed with SPSS version 19.0 software (SPSS, Inc, Chicago, Illinois, USA).

## RESULTS

### PATIENT CHARACTERISTICS

Of the 98 patients, 37(37.8%) were discharged, 43 (43.9%) died in hospital, and 18(18.4%) remained in hospital by March 29, 2020. The median age was 68.5 (63, 75) years, and 58.2% of the patients were female. The comorbidity was presented in most of the patients, with hypertension in 68.4%, diabetes in 17.3%, angina pectoris in 13.3%, previous myocardial infarction in 3.1%, previous PCI/CABG in 5.1%, chronic obstructive pulmonary disease in 8.2% and chronic renal dysfunction in 4.1% of the patients. Compared with survivors, non-survivors were more likely to have angina pectoris (20.9% vs 7.3%; *P*=0.048). It was notable that there were no significant differences in the age, sex, and other comorbidities including hypertension, previous PCI/CABG, chronic obstructive pulmonary disease and chronic renal dysfunction between the survivor and non-survivor group. (Table 1)

**Table 1.** Patient demographics.

| **Characteristics** | **Total**<br>**(n = 98)** | **Survivor**<br>**(n = 55)** | **Non-survivor**<br>**(n = 43)** | ***P* value** |
| --- | --- | --- | --- | --- |
| Age, years | 68.5 (63, 75) | 67 (63, 74) | 70 (65, 75) | 0.211 |
| Female | 57 (58.2%) | 27 (49.1%) | 30 (68.8%) | 0.039 |
| Alcohol consumption | 5 (5.1%) | 2 (3.6%) | 3 (7.0%) | 0.651 |
| Ever smoked | 11 (11.2%) | 6 (10.9%) | 5 (11.6%) | 1.000 |
| Hypertension | 67 (68.4%) | 39 (70.9%) | 28 (65.1%) | 0.541 |
| Diabetes mellitus | 17 (17.3%) | 10 (18.2%) | 7 (16.3%) | 0.805 |
| Angina pectoris | 13 (13.3%) | 4 (7.3%) | 9 (20.9%) | 0.048 |
| Previous myocardial infarction | 3 (3.1%) | 0 (0.0%) | 3 (7.0%) | 0.081 |
| Previous PCI/CABG | 5 (5.1%) | 1 (1.8%) | 4 (9.3%) | 0.165 |
| Chronic obstructive lung disease | 8 (8.2%) | 3 (5.5%) | 5 (11.6%) | 0.293 |
| Previous pneumonia | 6 (6.1%) | 2 (3.6%) | 4 (9.3%) | 0.400 |
| Malignant tumor | 5 (5.1%) | 4 (7.3%) | 1 (2.3%) | 0.381 |
| Chronic renal dysfunction | 4 (4.1%) | 2 (3.6%) | 2 (4.7%) | 1.000 |

PCI= percutaneous coronary intervention ; CABG=Coronary Artery Bypass Grafting

### LABORATORY FINDINGS

Most of 98 patients demonstrated an abnormal laboratory marker, with decreased lymphocyte percentage in 90.8%, elevated ALT, creatinine, D-dimer, myocardial injury markers and procalcitonin in 59.4%, 27.8%, 87.5%, 42.9% and 84.5% of the patients, respectively. Compared to the survivors, died patients showed lower lymphocyte percentage (100% vs 83.5%, *P*=0.015), higher proportion of elevated ALT (78.6% vs 44.4%, *P*=0.001), serum creatinine (sCr elevation of ≥50% or ≥0.3 mg/dl in hospital) (50.0% vs 10.9%, *P*<0.0001), cardiac injury marker (elevation of CK-MB or cardiac troponin I) (79.1% vs 14.5%, *P*=0.001). (Table 2)

**Table 2.** Patient Laboratory Findings.

| **Characteristics** | **Total**<br>**(n = 98)** | **Survivor**<br>**(n = 55)** | **Non-survivor**<br>**(n = 43)** | ***P* value** |
| --- | --- | --- | --- | --- |
| Lymphocyte percentage, % |  |  |  | 0.015 |
| <20 | 89 (90.8%) | 46 (83.6%) | 43 (100.0%) |  |
| ≥20 | 9 (9.2%) | 9 (16.4%) | 0 (0.0%) |  |
| ALT, IU/L |  |  |  | 0.001 |
| ≤50 | 39/96 (40.6%) | 30 (55.6%) | 9 (21.4%) |  |
| >50 | 57/96 (59.4%) | 24 (44.4%) | 33 (78.6%) |  |
| Creatinine, umol/L |  |  |  | 0.000 |
| ≤111 | 63 (64.3%) | 47 (85.5%) | 16 (37.2%) |  |
| >111 | 35 (35.7%) | 8 (14.5%) | 27 (62.8%) |  |
| Elevation of creatinine<br>(sCr)≥25% or ≥0.5<br>mg/dl I n hospital |  |  |  | 0.000 |
| No | 70/97 (72.7%) | 49(89.1%) | 21 (50.0%) |  |
| Yes | 27/97 (27.8%) | 6 (10.9%) | 21 (50.0%) |  |
| D-dimer, mg/L |  |  |  | 0.050 |
| ≤0.55 | 10/80 (12.5%) | 9 (20.0%) | 1 (2.9%) |  |
| >0.55 | 70/80 (87.5%) | 37 (80.0%) | 33 (97.1%) |  |
| Cardiac troponin I,<br>ng/ml |  |  |  | 0.000 |
| ≤0.04 | 27/59 (45.8%) | 21 (77.8%) | 6 (18.8%) |  |
| >0.04 | 32/59 (54.2%) | 6 (22.2%) | 26 (81.3%) |  |
| Creatine kinase<br>isoenzyme, IU/L |  |  |  | 0.000 |
| ≤24 | 61/97 (62.9%) | 49 (89.1%) | 12 (28.6%) |  |
| >24 | 36/97 (37.1%) | 6 (10.9%) | 30 (71.4%) |  |
| Cardiac injury marker<br>abnormality | 42 (42.9%) | 8 (14.5%) | 34 (79.1%) | 0.000 |
| Procalcitonin, ng/ml |  |  |  | 0.105 |
| ≤0.05 | 11/71 (15.5%) | 9 (23.1%) | 2 (6.3%) |  |
| >0.05 | 60/71 (84.5%) | 30 (76.9%) | 30 (93.8%) |  |

ALT = alanine aminotransferase. Cardiac injury marker abnormality = Cardiac troponin I >0.04 ng/ml or Creatine kinase isoenzyme >24 IU/L.

### TREATMENT AND COMPLICATIONS

The percentages of use of noninvasive ventilation and invasive mechanical ventilation were 52.0% (51patients) and 40.8% (40 patients). Of the 40 patients who received invasive mechanical ventilation, 33 patients (80.5%) died, 7 patients (12.3%) are still in hospital, and no patient was discharged. The proportion of antiviral therapy was the highest (86.7%), followed by antibiotics (80.6%), corticosteroids (65.3%) and traditional Chinese medicine treatment (28.6%). Only 3 patients (3.1%) received continuous renal replacement therapy, and 2 patients (2.0%) received ECMO therapy. Overall, the acute cardiac injury observed in 42.9% of the patients, followed by AKI (27.8%), and ARDS (26.5%). Other common complications included septic shock (3.1%), cardiogenic shock (2.0%), hypokalemia (1%) and hyperlactatemia (1%). Of note, the non-survivors demonstrated a higher rate of acute cardiac injury (79.1% vs 14.5%, *P*<0.0001), acute kidney injury (50.0%, vs 10.9%, *P*<0.0001) and ARDS (37.2 vs 18.2%, *P*=0.034) compared with survivors. (Table 3)

**Table 3.** Treatments and Complications.

| Characteristics | Total<br>(n = 98) | Survivor<br>(n = 55) | Non-survivor<br>(n = 43) | *P* value |
| --- | --- | --- | --- | --- |
| **Treatments** |  |  |  |  |
| Oxygen therapy |  |  |  |  |
| Noninvasive ventilator | 51 (52.0%) | 13 (23.6%) | 38 (88.4%) | 0.000 |
| Invasive mechanical ventilation | 40 (40.8%) | 5 (9.1%) | 35 (81.4%) | 0.000 |
| Tracheal cannula | 40 (40.8%) | 7 (12.3%) | 33 (80.5%) | 0.000 |
| Tracheotomy | 1 (1.0%) | 1 (1.8%) | 0 (0.0%) | 1.000 |
| ECMO | 2 (2.0%) | 1 (1.8%) | 1 (2.3%) | 1.000 |
| CRRT | 3 (3.1%) | 1 (1.8%) | 2 (4.7%) | 0.580 |
| Corticosteroids | 64 (65.3%) | 31 (56.4%) | 33 (76.7%) | 0.035 |
| Antiviral treatment | 85 (86.7%) | 51 (92.7%) | 34 (79.1%) | 0.048 |
| Traditional Chinese medicine | 28 (28.6%) | 27 (49.1%) | 1 (2.3%) | 0.000 |
| Antibiotics | 79 (80.6%) | 42 (77.2%) | 37 (85.4%) | 0.229 |
| **Complications** |  |  |  |  |
| Acute cardiac injury | 42 (42.9%) | 8 (14.5%) | 34 (79.1%) | 0.000 |
| Acute kidney injury | 27 (27.8%) | 6 (10.9%) | 21 (50.0%) | 0.000 |
| ARDS | 26 (26.5%) | 10 (18.2%) | 16 (37.2%) | 0.034 |
| Septic shock | 3 (3.1%) | 0 (0.0%) | 3 (7.0%) | 0.081 |
| Cardiogenic shock | 2 (2.0%) | 0 (0.0%) | 2 (4.7%) | 0.190 |
| Hypokalemia | 1 (1.0%) | 0 (0.0%) | 1 (2.3%) | 0.439 |
| Hyperlactatemia | 1 (1.0%) | 0 (0.0%) | 1 (2.3%) | 0.439 |
| Time from illness onset to high-flow nasal cannula oxygen therapy, days | 17.22±10.12 | 19.43±13.09 | 15.57±6.97 | 0.190 |
| Time from illness onset to noninvasive ventilator, days | 18.86±8.39 | 22.23±12.11 | 17.71±6.51 | 0.219 |
| Time from illness onset to tracheal cannula, days | 23.18±9.62 | 30.40±9.07 | 22.14±9.36 | 0.073 |
| Time from illness onset to the final follow-up date of endpoint, days | 37.60±17.19 | 45.58±16.09 | 27.40±12.61 | 0.000 |

### RISK FACTORS ASSOCIATED WITH IN-HOSPITAL DEATH

In univariable analysis, hazard ratio of in-hospital death was higher in patients with angina pectoris (HR 2.68, 1.28–5.58 *P*=0·009), previous myocardial infarction (HR 4.98, 1.48–16.77, *P*=0·009), elevated ALT (HR 2.48, 1.18–5.19 *P*=0·016), AKI (HR 3.31, 1.79-6.11 *P*<0·0001) and acute cardiac injury (HR 6.98, 3.34–14.59 *P*<0·0001). In the multivariable Cox regression model, we found that AKI (HR 2.60,1.15–5.86; *P*=0·021), acute cardiac injury (HR 6.57,1.89–22.79; *P*=0·005) and previous myocardial infarction (HR 6.33,1.70–23.65; *P*=0·006) were independently associated with increased hazard rate of death (Table 4). The mortality of the patients with acute cardiac injury (34 of 42 [81.0%] vs 9 of 56 [16.1%]; *P* < 0.001) or AKI (21 of 27 [77.8%] vs 21 of 70 [30.0%]; *P* < 0.001) is higher than those without acute cardiac injury or AKI. (Figure 1)

**Table 4.** Risk Factors Associated with in-hospital Death.

|  | Univariable |  | Multivariable |  |
| --- | --- | --- | --- | --- |
|  | HR (95%CI) | P value | HR (95%CI) | P value |
| Female (vs male) | 0.56 (0.29, 1.07) | 0.077 |  |  |
| Angina pectoris | 2.68 (1.28, 5.58) | 0.009 |  |  |
| Previous myocardial infarction | 4.98 (1.48, 16.77) | 0.009 | 6.33 (1.70, 23.65) | 0.006 |
| Lymphocyte percentage |  | 0.192 |  |  |
| <20 | 1 (ref) |  |  |  |
| ≥20 | 0.04 (0.00, 4.89) |  |  |  |
| ALT |  | 0.016 |  |  |
| ≤50 | 1 (ref) |  |  |  |
| >50 | 2.48 (1.18, 5.19) |  |  |  |
| Acute kidney injury | 3.31 (1.79, 6.11) | 0.000 | 2.60 (1.15, 5.86) | 0.021 |
| D-dimer |  | 0.116 |  |  |
| ≤0.55 | 1 (ref) |  |  |  |
| >0.55 | 4.92 (0.67, 35.93) |  |  |  |
| Acute cardiac injury | 6.98 (3.34, 14.59) | 0.000 | 6.57 (1.89, 22.79) | 0.005 |

ALT = alanine aminotransferase. Myocardial enzyme abnormality = Cardiac troponin I >0.04 or Creatine kinase isoenzyme >24.

**Figure 1.**
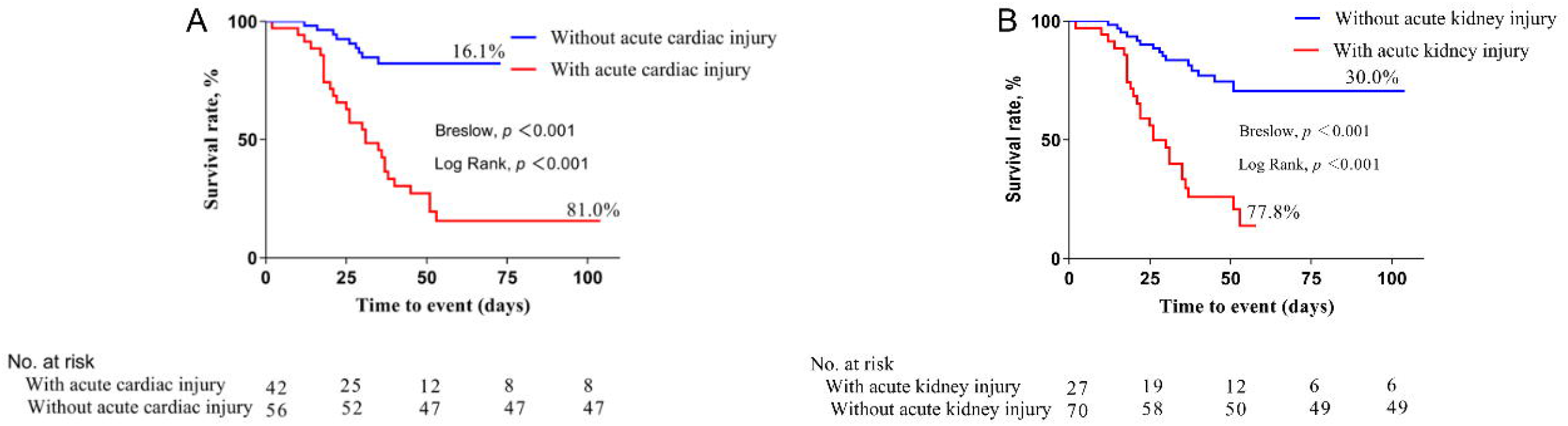
Mortality During Hospitalization Between Patients With vs Without Acute Cardiac Injury(A) or Acute Kidney Injury(B)

## DISCUSSION

Our present retrospective cohort study demonstrated a 43.9% of in-hospital death rate and identified several risk factors for in-hospital death in patients with critical COVID-19. Of a particular note, high incidence of acute cardiac injury and AKI was observed in our study population of patients with critical COVID-19, which are associated with a high risk of mortality during hospitalization.

By March 30, 2020, the number of COVID-19 had dramatically increased. According to the COVID-19 diagnosis and treatment program (trial sixth edition) issued by the National Health Commission of China(7), the patients were classified as clinically four types: mild, moderate, severe and critical type. For its rapidly progress in pulmonary injury and subsequently systemic complications(11), it is still the emphasis and difficulty in the treatment of critical type of COVID-19(12). At early stage of the COVID-19 outbreak, a descriptive study of 99 patients with moderate to critically ill presentation reported a 11% of mortality(5). A cohort study including 191 all hospitalized patients with a definite outcome (dead or discharged) reported a 28.3% of mortality(13). An observational study enrolled 21 critically ill patients from a public hospital in Washington State reported a 52.4% in-hospital death(1). A retrospective single-center study included 52 critically ill patients(3), and 32 (61·5%) patients had died at 28 days after hospitalization. In the present study, we enrolled 98 consecutive patients with critical COVID-19, and reported a 43.9% of in-hospital death rate. Taking into account of 18 patients still in hospital, we could infer that the mortality will continue to rise.

For the first time, we reported that AKI acted as an independent predictor for in-hospital death in COVID-19. Although studies reported that AKI occurred in COVID-19 patients, the association between AKI and risk of mortality has not be found yet. In the 1099 COVID-19 patients recently reported by Guan et al, the AKI incidence observed in unselected cases was 0.5% (14). A cohort study by Zhou et al in 191 patients reported an incidence of 15%(13). The difference in the prevalence rate of AKI may be attributed to the differences in enrolled patient populations. In the present study, the incidence of AKI is 27.8% in total, with an AKI incidence of 50% in the non-survivor group, which is consistent with Zhou’s study(13). Furthermore, even after adjusting for confounding factors, the multivariable adjusted Cox proportional hazard regression model still showed a significantly higher risk of death in patients with AKI than in those without AKI. The mechanism of coronavirus infection leading to AKI is not clear. According to existing studies(15,16), in addition to virus particles directly interacting with ACE2 receptor and overreacted immune responses, AKI in critical type of patients may also be caused by the following factors: hypotension, hypoxemia, electrolyte disorders, and massive use of immunosuppressive agents. However, more research is needed to clarify this issue.

Cardiac injury in COVID-19 has been reported in several studies(6,17,18). A report on 138 inpatients with COVID-19 showed that 7.2% of patients developed acute cardiac injury(4). A very recent study by Shi et al reported 19.7% incidence of acute cardiac injury in 416 unselected hospitalized patients with COVID-19^11^. An observational study enrolled 52 critically ill COVID patients demonstrated a 23% of cardiac injury(3). In the present, we reported a 42.9% of acute cardiac injury in patients with critical type of COVID-19, which is much higher than that in unselected or critical patients^10,11^. More importantly, in a Cox regression model, we found that patients with cardiac injury were associated with a higher hazard of death compared with those without cardiac injury, which is in line with Shi’s study^11^.

Previously, older age and D-dimer greater than 1 μg/L have been reported as important independent predictors of mortality in COVID-19 (13). However, in the present study, we do not find the predictive value of these factors for in-hospital death in critical patients. More importantly, there are no differences in age, sex and comorbidities including hypertension, diabetes and chronic obstructive pulmonary between the survivors and non-survivors in critical patients. Despite hypertension was not shown to be different between survivors and non-survivors, we should not neglect the fact that the percentage of patients with hypertension is extremely high (68.4%) in critical COVID-19 patients, while the data varied from 15% to 30% in total confirmed COVID-19 patients(6,13). Although with a small sample size, we found that previous myocardial infarction as an independent predictor of mortality. Most importantly, as discussed above we found acute cardiac injury increase the mortality in patients with COVID-19. Collectively, we presume that cardiovascular comorbidities and acute cardiac injury play a key role on in-hospital mortality in the critical COVID-19 patients.

Respiratory support is one of the most important treatment strategies for critical COVID-19 patients(19). In the early stage of the COVID-19 outbreak, a cohort study enrolled 191 patients conducted by Cao et al report that the invasive mechanical ventilation treatment is used in 32 patients (14%), of whom 31patients died with a 96.9% of mortality (13). In the present study, 40 patients (40.8%) receive invasive mechanical ventilation treatment, and 35 patients (87.5%) died, and 5 patients are still in hospital. It is notable that no patient undergoing mechanical ventilation treatment has been discharged so far. Our data confirm that patients in need of invasive mechanical ventilation appears a very high rate of in-hospital mortality. According to the existing data in different studies, ECMO is used in 1-3% of study population(4,13,20). But the survived case is rare. An observational study included 52 critical patients, of whom 6 received ECMO, and 5 died in hospital, with a mortality of 83.3%(3). In the present study, 2 patients received the ECMO treatment, of whom one died and the other one remained in-hospital. It is still controversial whether and when ECMO should be used in the critical patients with COVID-19. Therefore, the timing and management of invasive mechanical ventilation and ECMO need to be further optimized.

Our study has some limitations. Due to the retrospective study design, not all laboratory tests were done in all patients, including D-dimer and procalcitonin. Therefore, their role might be underestimated in predicting in-hospital death. Although we have adjusted for various variables that were associated with death in Cox regression analysis, there may be other potential confounders.

## CONCLUSIONS

A high incidence of acute cardiac injury and AKI were presented in COVID-19 classified as critical type, which were associated with high risk of in-hospital mortality.

## Data Availability

We would like to provide all data referred to the manuscript if they are requested.

## REFERENCE

1. Arentz M, Yim E, Klaff L et al. Characteristics and Outcomes of 21 Critically Ill Patients With COVID-19 in Washington State. JAMA 2020.

2. Qin C, Zhou L, Hu Z et al. Dysregulation of immune response in patients with COVID-19 in Wuhan, China. Clin Infect Dis 2020.

3. Yang X, Yu Y, Xu J et al. Clinical course and outcomes of critically ill patients with SARS-CoV-2 pneumonia in Wuhan, China: a single-centered, retrospective, observational study. Lancet Respir Med 2020.

4. Wang D, Hu B, Hu C et al. Clinical Characteristics of 138 Hospitalized Patients With 2019 Novel Coronavirus-Infected Pneumonia in Wuhan, China. JAMA 2020.

5. Chen N, Zhou M, Dong X et al. Epidemiological and clinical characteristics of 99 cases of 2019 novel coronavirus pneumonia in Wuhan, China: a descriptive study. Lancet 2020;395:507-513.

6. Huang C, Wang Y, Li X et al. Clinical features of patients infected with 2019 novel coronavirus in Wuhan, China. Lancet 2020;395:497-506.

7. WHO. Clinical management of severe acute respiratory infection when Novel coronavirus (nCoV) infection is suspected: interim guidance. Accessed January 31, 2020.:https://www.who.int/publications-detail/clinical-managementof-severe-acute-respiratory-infection-when-novelcoronavirus-(ncov)-infection-is-suspected.

8. New coronavirus pneumonia prevention and control program (6th ed.) (in Chinese). 2020: http://www.nhc.gov.cn/jkj/s3577/202003/4856d5b0458141fa9f376853224d41d7.shtml.

9. Ranieri VM, Rubenfeld GD, Thompson BT et al. Acute respiratory distress syndrome: the Berlin Definition. JAMA 2012;307:2526-33.

10. Stevens PE, Levin A, Members KDIGOCKDGDWG. Evaluation and management of chronic kidney disease: synopsis of the kidney disease: improving global outcomes 2012 clinical practice guideline. Ann Intern Med 2013;158:825-30.

11. Goh KJ, Choong MC, Cheong EH et al. Rapid Progression to Acute Respiratory Distress Syndrome: Review of Current Understanding of Critical Illness from COVID-19 Infection. Ann Acad Med Singapore 2020;49:1-9.

12. Zu ZY, Jiang MD, Xu PP et al. Coronavirus Disease 2019 (COVID-19): A Perspective from China. Radiology 2020:200490.

13. Zhou F, Yu T, Du R et al. Clinical course and risk factors for mortality of adult inpatients with COVID-19 in Wuhan, China: a retrospective cohort study. Lancet 2020;395: 1054-1062.

14. Wei-jie Guan Z-yN, Yu Hu, Wen-hua Liang, Chun-quan Ou, Jian-xing He, Lei Liu, Hong Shan, Chun-liang Lei, David SC Hui, Bin Du, Lan-juan Li, Guang Zeng, Kowk-Yung Yuen, Ru-chong Chen, Chun-li Tang, Tao Wang, Ping-yan Chen, Jie Xiang, Shi-yue Li, Jin-lin Wang, Zi-jing Liang, Yi-xiang Peng, Li Wei, Yong Liu, Ya-hua Hu, Peng Peng, Jian-ming Wang, Ji-yang Liu, Zhong Chen, Gang Li, Zhi-jian Zheng, Shao-qin Qiu, Jie Luo, Chang-jiang Ye, Shao-yong Zhu, Nan-shan Zhong. Clinical characteristics of 2019 novel coronavirus infection in China. medRxiv 2020;https://doi.org/10.1101/2020.02.06.20020974.

15. Zhou P, Yang XL, Wang XG et al. A pneumonia outbreak associated with a new coronavirus of probable bat origin. Nature 2020;579:270-273.

16. Rodriguez-Morales AJ, Cardona-Ospina JA, Gutierrez-Ocampo E et al. Clinical, laboratory and imaging features of COVID-19: A systematic review and meta-analysis. Travel Med Infect Dis 2020:101623.

17. Chen C, Yan JT, Zhou N, Zhao JP, Wang DW. [Analysis of myocardial injury in patients with COVID-19 and association between concomitant cardiovascular diseases and severity of COVID-19]. Zhonghua Xin Xue Guan Bing Za Zhi 2020;48:E008.

18. Shi S, Qin M, Shen B et al. Association of Cardiac Injury With Mortality in Hospitalized Patients With COVID-19 in Wuhan, China. JAMA Cardiol 2020.

19. Physician CccoCAoC, Society RaccgoCT, Society RcgoCT. [Conventional respiratory support therapy for Severe Acute Respiratory Infections (SARI): Clinical indications and nosocomial infection prevention and control]. Zhonghua Jie He He Hu Xi Za Zhi 2020;43:189-194.

20. Ramanathan K, Antognini D, Combes A et al. Planning and provision of ECMO services for severe ARDS during the COVID-19 pandemic and other outbreaks of emerging infectious diseases. Lancet Respir Med 2020.

